# Covid-19 Vaccination in Pregnancy: A Systematic Review

**DOI:** 10.1101/2021.07.04.21259985

**Authors:** Nando Reza Pratama, Ifan Ali Wafa, David Setyo Budi, Manesha Putra, Manggala Pasca Wardhana, Citrawati Dyah Kencono Wungu

## Abstract

**Objective:** Pregnancy is a risk factor for severe Covid-19. Looking for safe vaccines that evoke protective maternal and fetal antibody response is important.

**Methods:** We searched from registries (ClinicalTrials.gov, the WHO Clinical Trial Registry, and the EU Clinical Trial Registry) and databases (MEDLINE, ScienceDirect, Cochrane Library, Proquest, and Springer) up until June 20, 2021. Articles were selected based on inclusion and exclusion criteria after duplicates were removed. Infection rate, maternal antibody response, placental antibody transfer, and adverse events were described. This systematic review was performed with quality assessment and semi-quantitative synthesis according to PRISMA guidelines.

**Results:** Twelve observational studies with a total of 40.509 pregnant women included. The mRNA based vaccines (Pfizer-BioNTech and Moderna) can prevent future SARS-CoV-2 infections (p=0.0004). Both vaccines did not affect pregnancy, delivery, and neonatal outcomes. The most commonly encountered adverse reactions are injection-site pain, fatigue, and headache but only transient. Antibody responses were rapid after the prime dose of vaccines. After booster, antibody responses were higher and associated with better placental antibody transfer. Longer intervals between first vaccination dose and delivery were also associated with higher antibody fetal IgG and better antibody transfer ratio.

**Conclusions:** The Pfizer-BioNTech and Moderna vaccines are efficacious for preventing future SARS-CoV-2 infections. These vaccines can be considered as a safe option for pregnancy and their fetus. Two doses of vaccines were recommended for more robust maternal and fetal antibody responses. Longer latency was associated with higher fetal antibody responses.

**Systematic Review Registration:** PROSPERO (CRD42021261684)

## INTRODUCTION

The impact of coronavirus disease 2019 (Covid-19) on pregnancy remains unclear due to insufficient information, while most available studies have evaluated the impact of the disease only in the general population. The risk of severe Covid-19 in pregnancy appears to be no greater than for the general population; pregnant women are at a higher risk for acquiring viral respiratory infections and severe pneumonia due to the unique physiological changes in their immune and cardiopulmonary systems.^1,2^ Although most pregnant women had mild to moderate symptoms only, SARS-CoV-2 infection is found more severe in pregnant women than their nonpregnant counterparts, with an increased risk of hospital admission, intensive care unit stay, and death.^3^

Despite their higher risk of SARS-CoV-2 infection, pregnant and lactating women were not included in any initial Covid-19 vaccine trials, resulting in data scarcity to guide vaccine decision making in these populations.^4^ A prior study revealed that most pregnant women with Covid-19 admitted to the hospital were asymptomatic, which allows these undetected patients to transmit the virus to others.^5,6^ This finding shows that efforts to prevent SARS-CoV-2 infection, one of them by vaccination, are critical for investigation on this population.

To facilitate comprehending the Covid-19 vaccine in pregnancy, we performed a systematic review to critically evaluate and summarize the latest evidence of Covid-19 vaccination regarding the efficacy, immunogenicity, and safety profile in the pregnant population.

## METHODS

This systematic review adhered to PRISMA (Preferred Reporting Items for Systematic Review and Meta-Analysis) 2020 guidelines^7^ and has been previously registered in the PROSPERO database (CRD42021261684).

### Eligibility criteria

We accepted any study (retrospective, prospective, cohort, randomized controlled trials (RCT), case series, case control, cross-sectional, crossover) to be included in the review. The authors screened the title and abstract of independently for eligible studies based on the following criteria: (1) pregnant women; (2) adult (≥18 years) female study population; (3) the study involved Covid-19 vaccine of interest; (4) the study compared the intervention group with control (non-pregnant women, unvaccinated, or none); (5) eligible studies should have reported at least one of our outcomes of interest; (6) English language. Our primary outcomes included infection rate, maternal titer antibody, local and systemic adverse events. Our secondary outcomes included neonatal outcome, cord blood titer antibody, and placental transfer ratio. We excluded review articles, irrelevant, non-human studies, and duplications.

### Search strategy and selection of studies

The authors comprehensively conducted keyword searching of articles published in trial registries (ClinicalTrials.gov, the WHO Clinical Trial Registry, and the EU Clinical Trial Registry) and databases (MEDLINE, ScienceDirect, Cochrane Library, Proquest, and Springer) up until June 20, 2021. Our research is limited to studies involving humans with no language restrictions. Manual search, including in BioRxiv and MedRxiv, and the bibliographical search were also conducted to obtain additional evidence. The following keywords were used: “((SARS-CoV-2) OR (COVID-19)) AND ((pregnancy) OR (pregnant)) AND ((vaccine) OR (vaccination))”. The search strategies are available in ***Supplementary Materials***. We exported all studies retrieved from the electronic searches into Mendeley reference manager for removing duplication and screening. The two review authors (NRP and IAW) independently screened the title and abstract of the articles to identify potentially eligible studies and subsequently screened the full texts independently. Any disagreements between the two review authors were resolved by discussion until reaching consensus. Excluded studies were described in the PRISMA flow diagram alongside their reasons for exclusion (**Figure 1**).

**Figure 1.**
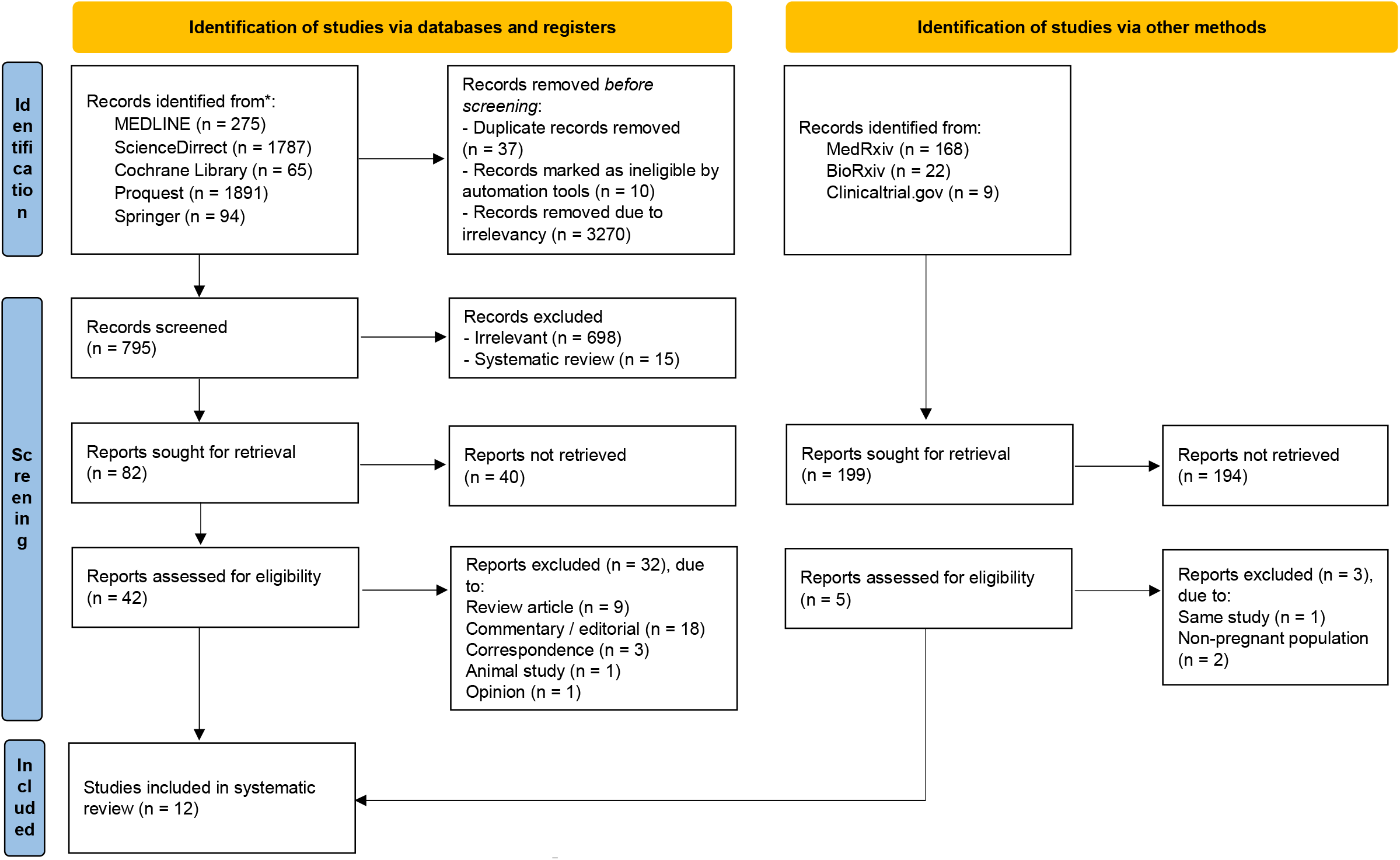
PRISMA flow diagram of study selection process^7^

### Data extraction

The review authors (NRP, IAW, and DSB) independently extracted relevant data using structured and standardized form from each selected study. The following information was extracted: first author’s name and publication year, study design, country, sample size, gestational age at first vaccine, sample size, age, sample collection, vaccine type, and outcomes (infection rate, maternal titer antibody, cord blood titer antibody, placental transfer ratio and local and systemic adverse events). Any disagreements between the review authors were consulted with the expert and resolved by discussion until reaching consensus.

### Quality assessment

Two review authors (IAW and DSB) independently assessed the risk of bias from each included studies using Newcastle-Ottawa Scale (NOS) assessment tool for cohort studies and Joanna Briggs Institute (JBI) critical appraisal checklist for case report, case series, and cross-sectional studies.^8,9^ Newcastle-Ottawa Scale contains eight items within three domains, including patient selection, comparability, and outcomes. A study with scores 7-9, 4-6, and 0-3 were considered to be high, moderate, and low-quality, respectively. Any discrepancies were resolved by discussion until reaching consensus.

### Statistical analysis

Due to important differences in comparison of each study and various outcome measures, we could not generate meta-analyses of the included studies but rather we performed narratively synthesized the evidence using Synthesis Without Meta-analysis (SWiM) reporting guideline (intended to complement the PRISMA guidelines in these cases).

## RESULTS

### Study selection

The search strategy yielded 4112 and 199 records, respectively, from database searching and additional searching, in medRxiv, bioRxiv, and clinicaltrial.gov. After screening the titles and abstracts, 47 potentially eligible articles were reviewed. After the full-text assessment, twelve studies were included in this systematic review. The process of study selection in this review is described in the PRISMA flow diagram (**Figure 1**), along with the reason for exclusion.

### Quality assessment

Eight cohort studies were assessed using the NOS assessment tool and considered as high quality since all of the studies scored 7-9 (see ***Table S1 in Supplementary Materials***). The quality assessment of case report, case series, and cross-sectional studies using the JBI critical appraisal checklist was summarized in Supplementary Materials (**Table S2 – S4**).

### Study characteristics

There are twelve observational studies with a total of 40.509 pregnant women who received Covid-19 vaccines included in this systematic review. Among these studies, ten studies (six cohort, one cross-sectional, one case series, and two case report) were conducted in the United States of America (USA) while other two cohort studies were carried out in Israel in 2021. The maternal age ranged from 16 to 54 years. All pregnant women reported receiving mRNA vaccine, either Pfizer–BioNTech or Moderna vaccine, except for four pregnant women who received unknown types of vaccine.^18^ Some studies compared vaccinated pregnant women with unvaccinated pregnant women, either naturally infected or not infected, or vaccinated non-pregnant women. The efficacy outcome was desribed as infection rate, while immunogenicity was described as maternal antibody response, fetal antibody response, and transplacental antibody transfer. Safety outcome was described as the adverse events, maternal outcomes, and neonatal outcomes. Adverse events were divided into local and systemic, local adverse events include injection-site pain and soreness, while systemic adverse events include fatigue, headache, myalgia, chills, fever and nausea.

**Table 1A.**
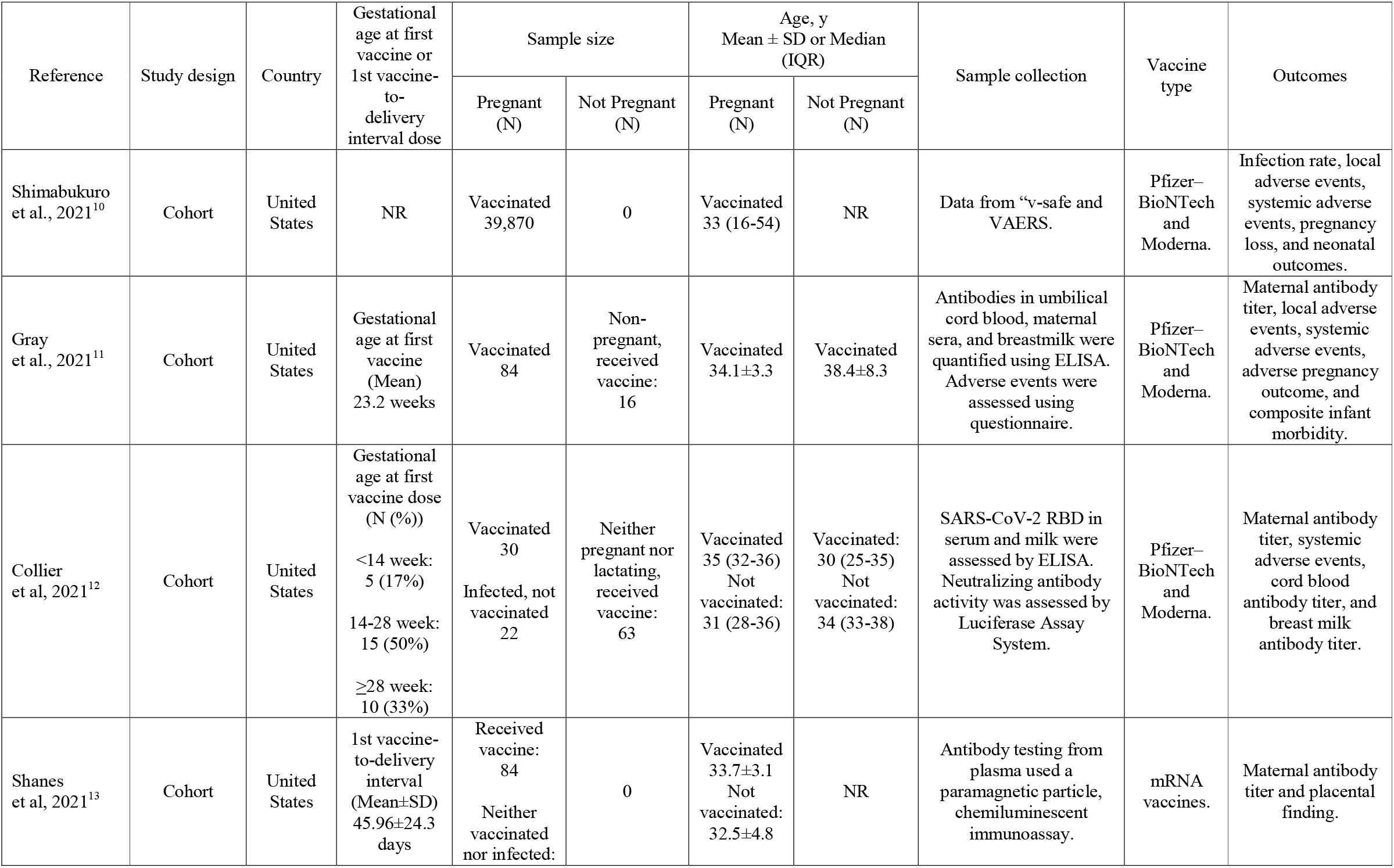
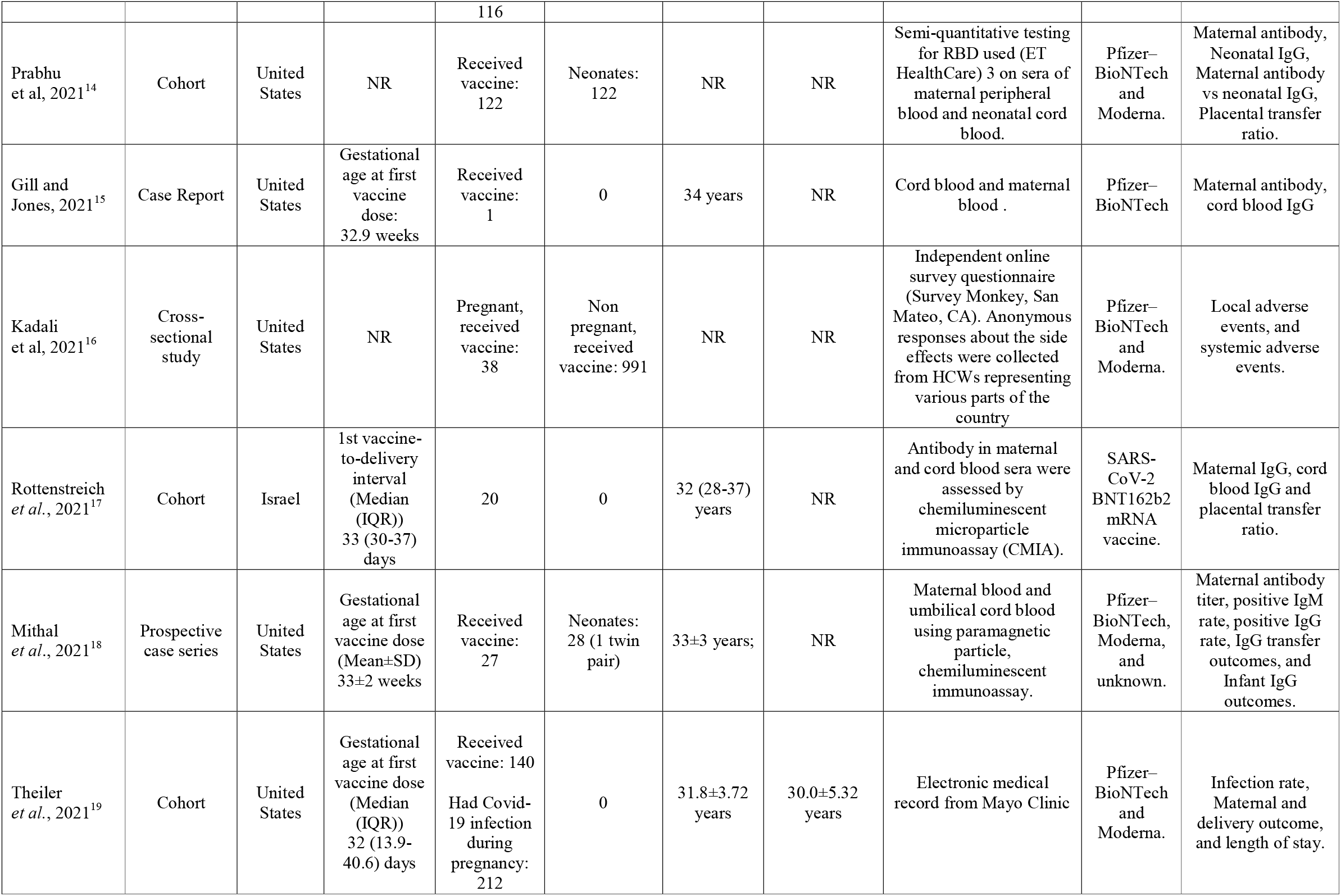
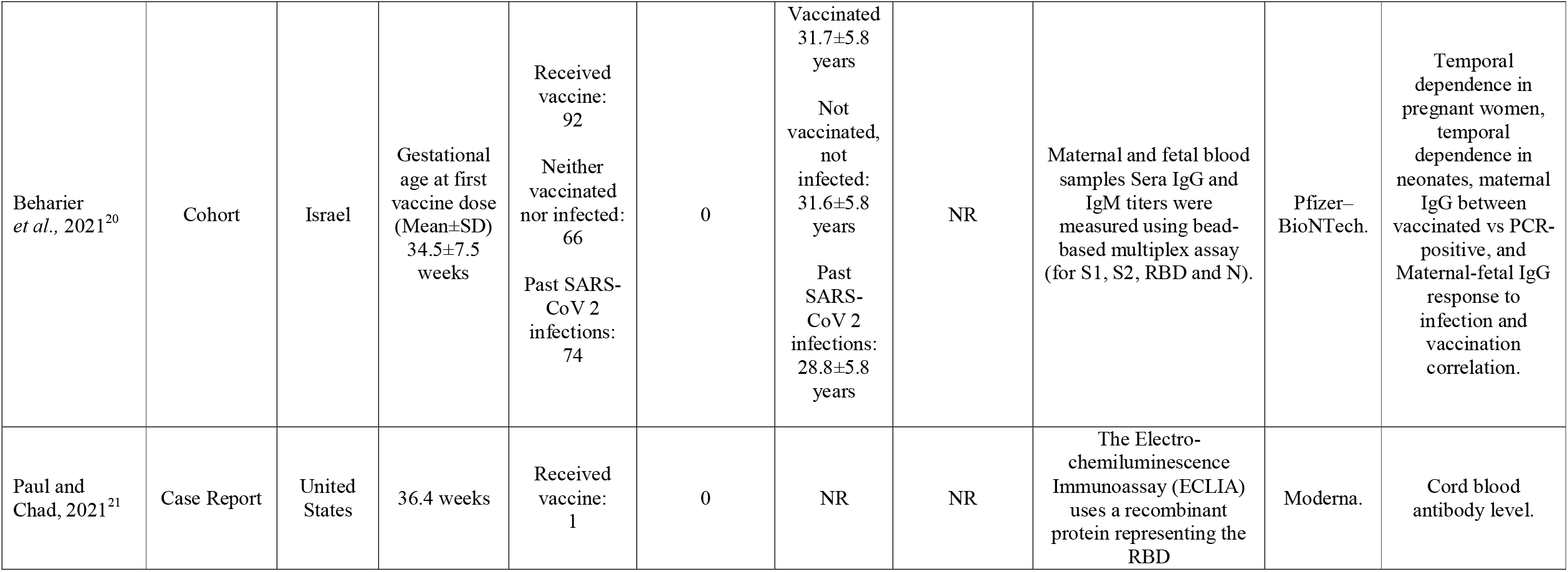
Characteristics of the included studies

| Reference | Study design | Country | Gestational age at first vaccine or 1st vaccine-to-delivery interval dose | Sample size | | Age, y<br>Mean ± SD or Median (IQR) | | Sample collection | Vaccine type | Outcomes |
| --- | --- | --- | --- | --- | --- | --- | --- | --- | --- | --- |
|  |  |  |  | Pregnant (N) | Not Pregnant (N) | Pregnant (N) | Not Pregnant (N) |  |  |  |
| Shimabukuro et al., 2021<sup>10</sup> | Cohort | United States | NR | Vaccinated 39,870 | 0 | Vaccinated 33 (16-54) | NR | Data from "v-safe and VAERS. | Pfizer–BioNTech and Moderna. | Infection rate, local adverse events, systemic adverse events, pregnancy loss, and neonatal outcomes. |
| Gray et al., 2021<sup>11</sup> | Cohort | United States | Gestational age at first vaccine (Mean) 23.2 weeks | Vaccinated 84 | Non-pregnant, received vaccine: 16 | Vaccinated 34.1±3.3 | Vaccinated 38.4±8.3 | Antibodies in umbilical cord blood, maternal sera, and breastmilk were quantified using ELISA. Adverse events were assessed using questionnaire. | Pfizer–BioNTech and Moderna. | Maternal antibody titer, local adverse events, systemic adverse events, adverse pregnancy outcome, and composite infant morbidity. |
| Collier et al., 2021<sup>12</sup> | Cohort | United States | Gestational age at first vaccine dose (N (%))<br><14 week: 5 (17%)<br>14-28 week: 15 (50%)<br>≥28 week: 10 (33%) | Vaccinated 30<br>Infected, not vaccinated 22 | Neither pregnant nor lactating, received vaccine: 63 | Vaccinated 35 (32-36)<br>Not vaccinated: 31 (28-36) | Vaccinated: 30 (25-35)<br>Not vaccinated: 34 (33-38) | SARS-CoV-2 RBD in serum and milk were assessed by ELISA. Neutralizing antibody activity was assessed by Luciferase Assay System. | Pfizer–BioNTech and Moderna. | Maternal antibody titer, systemic adverse events, cord blood antibody titer, and breast milk antibody titer. |
| Shanes et al., 2021<sup>13</sup> | Cohort | United States | 1st vaccine-to-delivery interval (Mean±SD) 45.96±24.3 days | Received vaccine: 84<br>Neither vaccinated nor infected: | 0 | Vaccinated 33.7±3.1<br>Not vaccinated: 32.5±4.8 | NR | Antibody testing from plasma used a paramagnetic particle, chemiluminescent immunoassay. | mRNA vaccines. | Maternal antibody titer and placental finding. |
|  |  |  |  | 116 |  |  |  |  |  |  |
| Prabhu et al, 2021<sup>14</sup> | Cohort | United States | NR | Received vaccine: 122 | Neonates: 122 | NR | NR | Semi-quantitative testing for RBD used (ET HealthCare) 3 on sera of maternal peripheral blood and neonatal cord blood. | Pfizer–BioNTech and Moderna. | Maternal antibody, Neonatal IgG, Maternal antibody vs neonatal IgG, Placental transfer ratio. |
| Gill and Jones, 2021<sup>15</sup> | Case Report | United States | Gestational age at first vaccine dose: 32.9 weeks | Received vaccine: 1 | 0 | 34 years | NR | Cord blood and maternal blood . | Pfizer–BioNTech | Maternal antibody, cord blood IgG |
| Kadali et al, 2021<sup>16</sup> | Cross-sectional study | United States | NR | Pregnant, received vaccine: 38 | Non pregnant, received vaccine: 991 | NR | NR | Independent online survey questionnaire (Survey Monkey, San Mateo, CA). Anonymous responses about the side effects were collected from HCWs representing various parts of the country | Pfizer–BioNTech and Moderna. | Local adverse events, and systemic adverse events. |
| Rottenstreich et al., 2021<sup>17</sup> | Cohort | Israel | 1st vaccine-to-delivery interval (Median (IQR)) 33 (30-37) days | 20 | 0 | 32 (28-37) years | NR | Antibody in maternal and cord blood sera were assessed by chemiluminescent microparticle immunoassay (CMIA). | SARS-CoV-2 BNT162b2 mRNA vaccine. | Maternal IgG, cord blood IgG and placental transfer ratio. |
| Mithal et al., 2021<sup>18</sup> | Prospective case series | United States | Gestational age at first vaccine dose (Mean±SD) 33±2 weeks | Received vaccine: 27 | Neonates: 28 (1 twin pair) | 33±3 years; | NR | Maternal blood and umbilical cord blood using paramagnetic particle, chemiluminescent immunoassay. | Pfizer–BioNTech, Moderna, and unknown. | Maternal antibody titer, positive IgM rate, positive IgG rate, IgG transfer outcomes, and Infant IgG outcomes. |
| Theiler et al., 2021<sup>19</sup> | Cohort | United States | Gestational age at first vaccine dose (Median (IQR)) 32 (13.9-40.6) days | Received vaccine: 140<br>Had Covid-19 infection during pregnancy: 212 | 0 | 31.8±3.72 years | 30.0±5.32 years | Electronic medical record from Mayo Clinic | Pfizer–BioNTech and Moderna. | Infection rate, Maternal and delivery outcome, and length of stay. |
| Beharier<br>*et al.*, 2021<sup>20</sup> | Cohort | Israel | Gestational<br>age at first<br>vaccine dose<br>(Mean±SD)<br>34.5±7.5<br>weeks | Received<br>vaccine:<br>92<br>Neither<br>vaccinated<br>nor infected:<br>66<br>Past SARS-CoV 2<br>infections:<br>74 | 0 | Vaccinated<br>31.7±5.8<br>years<br>Not<br>vaccinated,<br>not<br>infected:<br>31.6±5.8<br>years<br>Past<br>SARS-CoV 2<br>infections:<br>28.8±5.8<br>years | NR | Maternal and fetal blood<br>samples Sera IgG and<br>IgM titers were<br>measured using bead-<br>based multiplex assay<br>(for S1, S2, RBD and N). | Pfizer–<br>BioNTech. | Temporal<br>dependence in<br>pregnant women,<br>temporal<br>dependence in<br>neonates, maternal<br>IgG between<br>vaccinated vs PCR-positive, and<br>Maternal-fetal IgG<br>response to<br>infection and<br>vaccination<br>correlation. |
| Paul and<br>Chad, 2021<sup>21</sup> | Case Report | United<br>States | 36.4 weeks | Received<br>vaccine:<br>1 | 0 | NR | NR | The Electro-<br>chemiluminescence<br>Immunoassay (ECLIA)<br>uses a recombinant<br>protein representing the<br>RBD | Moderna. | Cord blood<br>antibody level. |

**Table 1B.** Outcomes of the individual studies

| Reference | Infection rate<br>N (%) | | Maternal SARS-CoV 2 antibody titer<br>(Mean ± SD or Median (IQR)) | | Local adverse event<br>N (%) | |
| --- | --- | --- | --- | --- | --- | --- |
|  | Intervention | Comparison | Pregnant | Non-Pregnant | Pregnant | Non-Pregnant |
| Shimabukuro *et al.*, 2021<sup>10</sup> | **Pfizer–BioNTech vaccine**<br>$\leq 14$ days after first eligible dose of vaccination: 3 (0.1%)<br>$> 14$ days after first eligible dose of vaccination: 9 (0.4%) | **Moderna vaccine**<br>$\leq 14$ days after first eligible dose of vaccination: 7 (0.4%)<br>$> 14$ days after first eligible dose of vaccination: 3 (0.2%) | NR | NR | **Pfizer–BioNTech vaccine (1<sup>st</sup> dose vs 2<sup>nd</sup> dose)**<br>Injection-site pain: 7602 (84%) vs 5886 (89%)<br>Injection-site redness: 160 (2%) vs 169 (3%)<br>Injection-side itching: 103 (1%) vs 109 (2%)<br>**Moderna vaccine (1<sup>st</sup> vs 2<sup>nd</sup> dose)**<br>Injection-site pain: 7360 (93%) vs 5388 (96%)<br>Injection-site redness: 348 (4%) vs 491 (9%)<br>Injection-side itching: 157 (2%) vs 193 (3%) | NR |
| Gray *et al.*, 2021<sup>11</sup> | NR | NR | NR | NR | **First dose vaccine**<br>Injection-site soreness: 73 (88%)<br>Injection site reaction or rash: 1 (1%)<br>**Second dose vaccine**<br>Injection-site soreness: 44 (57%)<br>Injection site reaction or rash: 1 (1%) | **First dose vaccine**<br>Injection-site soreness: 12 (75%)<br>Injection site reaction or rash: 0 (0%)<br>**Second dose vaccine**<br>Injection-site soreness: 12 (75%)<br>Injection site reaction or rash: 0 (0%) |
| Collier *et al.*, 2021<sup>12</sup> | NR | NR | **Vaccinated**<br>RBD IgG (median): 27,601 AU<br>Neutralizing Ab (median): 910 AU<br>**Infected**<br>RBD IgG (median): 1,321 AU<br>Neutralizing Ab (median): 148 AU | **Vaccinated**<br>RBD IgG (median): 37,839 AU<br>Neutralizing Ab (median): 901 AU<br>**Infected**<br>RBD IgG (median): 771 AU<br>Neutralizing Ab (median): 193 AU | NR | NR |
| Shanes *et al.*, 2021<sup>13</sup> | NR | NR | **Vaccinated**<br>RBD IgG: 22.8±14.5<br>RBD IgM: 4.1±13.2<br>**Unvaccinated**<br>RBD IgG: 0.04±0.05 | NR | NR | NR |
|  |  |  | RBD IgM: 0.19±0.12 |  |  |  |
| Prabhu *et al.*, 2021<sup>14</sup> | NR | NR | NR | NR | NR | NR |
| Gill and Jones, 2021<sup>15</sup> | NR | NR | SARS-CoV-2 IgG titer: 1:25600 (+) | NR | NR | NR |
| Kadali *et al.*, 2021<sup>16</sup> | NR | NR | NR | NR | Sore arm or pain: 37 (97%)<br>Itching: 2 (5%)<br>Muscle spasm: 1 (3%) | Sore arm or pain: 894 (90%)<br>Itching: 98 (10%)<br>Muscle spasm: 103 (10%) |
| Rottenstreich *et al.*, 2021<sup>17</sup> | NR | NR | Anti-S IgG: 319 (211-1033) AU/mL<br>Anti-RBD-Specific IgG: 11,150 (6154-17,575) AU/mL | NR | NR | NR |
| Mithal *et al.*, 2021<sup>18</sup> | NR | NR | NR | NR | NR | NR |
| Theiler *et al.*, 2021<sup>19</sup> | **Vaccinated:**<br>None: 138 (99%)<br>Trimester 1: 0 (0%)<br>Trimester 2: 2 (1%)<br>Trimester 3: 0 (0%) | **Not vaccinated:**<br>None: 1652 (89%)<br>Trimester 1: 26 (1%)<br>Trimester 2: 84 (5%)<br>Trimester 3: 100 (5%) | NR | NR | NR | NR |
|  | p=0.0004 |  |  |  |  |  |
| Beharier *et al.*, 2021<sup>20</sup> | NR | NR | NR | NR | NR | NR |
| Paul and Chad, 2021<sup>21</sup> | NR | NR | NR | NR | NR | NR |

| Reference | Systemic adverse events N (%) |  | Others |
| --- | --- | --- | --- |
|  | Pregnant | Non-Pregnant |  |
| Shimabukuro et al, 2021<sup>10</sup> | **Pfizer–BioNTech vaccine (1<sup>st</sup> dose vs 2<sup>nd</sup> dose)**<br>Fatigue: 2406 (27%) vs 4231 (64%)<br>Headache: 1497 (17%) vs 3138 (47%)<br>Myalgia: 795 (9%) vs 2916 (44%)<br>Chills: 254 (3%) vs 1747 (26%)<br>Fever or felt feverish: 256 (3%) vs 1648 (25%)<br>Measured temperature $\geq 38^{\circ}\text{C}$: 30 (0%) vs 315 (5%)<br>Nausea: 492 (5%) vs 1356 (20%)<br>**Moderna vaccine (1<sup>st</sup> vs 2<sup>nd</sup> dose)**<br>Fatigue: 2616 (33%) vs 4541 (81%)<br>Headache: 1581 (20%) vs 3662 (65%)<br>Myalgia: 1167 (15%) vs 3722 (66%)<br>Chills: 442 (6%) vs 2755 (49%)<br>Fever or felt feverish: 453 (6%) vs 2594 (46%)<br>Measured temperature $\geq 38^{\circ}\text{C}$: 62 (1%) vs 664 (12%)<br>Nausea: 638 (8%) vs 1909 (34%) | NR | **Maternal and delivery outcomes**<br>Pregnancy loss among complete pregnancy (N (%))<br>- Abortion: 104 (12.6%)<br>- Stillbirth: 1 (0.1%)<br>Neonatal outcome among live-born infants (N (%))<br>- Preterm birth: 60 (9.4%)<br>- Small size for gestational age: 23 (3.2%)<br>- Congenital anomalies (N=16; 2.2%)<br>- Neonatal death (N=0; 0%) |
| Gray et al, 2021<sup>11</sup> | **First dose vaccine**<br>Headache: 7 (8%)<br>Muscle aches: 2 (2%)<br>Fatigue: 12 (14%)<br>Fever or chills: 1 (1%)<br>**Second dose vaccine**<br>Headache: 25 (32%)<br>Muscle aches: 37 (48%)<br>Fatigue: 41 (53%) | **First dose vaccine (N (%))**<br>Headache: 5 (31%)<br>Muscle aches: 2 (12%)<br>Fatigue: 6 (38%)<br>Fever or chills: 1 (6%)<br>**Second dose vaccine (N (%))**<br>Headache: 6 (38%)<br>Muscle aches: 7 (44%)<br>Fatigue: 9 (56%) | **Adverse pregnancy outcome (N (%))**<br>Fetal growth restriction: 0 (0%)<br>Preeclampsia/gestational hypertension: 0 (0%)<br>Preterm delivery (spontaneous): 1 (8%)<br>Preterm delivery (medically indicated): 0 (0%)<br>**Composite infant morbidity (N (%))**<br>Supplemental oxygen/CPAP: 1 (8%)<br>TTN: 1 (8%)<br>Special care nursery admission: 0 (0%) |
|  | Fever or chills: 25 (32%) | Fever or chills: 8 (50%) | NICU admission: 2 (15%)<br>Respiratory distress syndrome: 0 (0%)<br>Necrotizing enterocolitis: 0 (0%)<br>Sepsis: 0 (0%)<br>Assisted ventilation: 0 (0%)<br>Seizure: 0 (0%)<br>Grade 3/4 intraventricular hemorrhage: 0 (0%)<br>Death: 0 (0%)<br>**IgG Spike response**<br>Pregnant V1 vs Pregnant V0: P<0.0001<br>Pregnant V2 vs Pregnant V0: P<0.0001<br>Pregnant V2 vs Pregnant V1: P<0.05<br>Cord blood IgG titer vs time from maternal V2 corr.(r): 0.8; p=0.01<br>Vaccinated pregnant vs natural infection pregnant titer: P<0.0001<br>**IgG RBD response**<br>Pregnant V1 vs Pregnant V0: P<0.01<br>Pregnant V2 vs Pregnant V0: P<0.0001<br>Pregnant V2 vs Pregnant V1: P<0.001<br>Cord blood IgG titer vs time from maternal V2 corr.(r): 0.50; p=0.17<br>**Neutralizing antibody titer (umbilical cord vs maternal serum)**<br>Medial (IQR): 104 (61.2-188.2) vs 52.3 (11.7-69.6); P=0.05<br>**Antibodies transfer from maternal to cord blood**<br>Spike IgG3 (r): 0.93; p=0.03<br>RBD IgG3 (r): 0.81; p=0.07 |
| Collier et al, 2021<sup>12</sup> | Fever after first dose: 0 (0%)<br>Fever after second dose: 4 (14%) | Fever after first dose: 1 (2%)<br>Fever after second dose: 27 (52%) | **RBD IgG titer median (mother serum vs cord blood)**<br>Vaccinated: 14953 vs 19873<br>Infected: 1324 vs 635<br>**Neutralizing antibodies titer median (mother serum vs cord blood)**<br>Vaccinated: 1016 AU vs 324<br>Infected: 151 vs 164<br>**RBD IgG against SARS-CoV-2 Variants of Concern**<br>Binding antibody responses were comparable against wild type USA-WA1/2020 and B.1.1.7 RBD proteins in nonpregnant, pregnant, and lactating women and in infant cord samples but were lower for the B.1.351 RBD protein. |
| Shanes et al, 2021<sup>13</sup> | NR | NR | NR |
| Prabhu et al, 2021<sup>14</sup> | NR | NR | **Positive maternal antibody rate**<br>Women with detectable:<br>- IgG & IgM (N (%)): 87 (71%)<br>- IgG only (N (%)): 19 (16%)<br>Women with no detectable IgG & IgM (N (%)): 16 (13%)<br>**Positive neonatal antibody rate**<br>IgG from whom the mother received:<br>- One vaccine dose (N (%)): 24 (43.6%)<br>- Two vaccine doses (N (%)): 65 (98.5%)<br>**Placental transfer outcome**<br>Maternal IgG and neonatal IgG correlation (R): 0.89, p<2.2 e-16<br>Placental transfer ratio and weeks elapsed since maternal vaccination dose 2 correlation (R): 0.8, p=2.6 e-16 |
| Gill and Jones, 2021<sup>15</sup> | NR | NR | **Cord blood antibody:**<br>SARS-CoV-2 IgG titer: 1:25600 (+) |
| Kadali et al., | **Pregnant** | **Non pregnant** | NR |
| 2021<sup>16</sup> | Fatigue: 22 (58%)<br>Headache: 19 (50%)<br>Chills: 18 (47%)<br>Myalgia: 13 (34%)<br>Nausea: 11 (29%)<br>Fever: 6 (16%)<br>Seizure*: 1 (3%) | Fatigue: 643 (65%)<br>Headache: 519 (52%)<br>Chills: 424 (43%)<br>Myalgia: 488 (49%)<br>Nausea: 211 (21%)<br>Fever: 279 (28%)<br>Seizure*: 0 (0%) |  |
| Rottenstreich<br>*et al.*, 2021<sup>17</sup> | NR | NR | **Cord-Blood level**<br>Anti-S IgG (median (IQR)): 193 (111-260) AU/mL<br>Anti-RBD-specific IgG (median (IQR)): 3494 (1817-6163) AU/mL<br>**Placental transfer ratio**<br>Anti-S IgG (median (IQR)): 0.44 (0.25-0.61)<br>Anti-RBD-specific IgG (median (IQR)): 0.34 (0.27-0.56) |
| Mithal *et al.*,<br>2021<sup>18</sup> | NR | NR | **Positive IgM rate**<br>Maternal serum (N (%)): 15 (56%)<br>Cord blood (N (%)): 0 (0%)<br>**Positive IgG rate**<br>Maternal serum (N (%)): 26 (96%)<br>Cord blood (N (%)): 25 (89%)<br>**IgG transfer outcomes**<br>Maternal to infant (mean±SD): 1.0±0.6<br>Latency from vaccination to delivery vs IgG transfer ratio correlation (β): 0.2 (95%CI 0.1-0.2)<br>**Infant IgG outcomes**<br>Having received the 2<sup>nd</sup> vaccine dose vs infant IgG level correlation (β): 19.0 (95%CI 7.1-30.8)<br>Latency from vaccination to delivery vs infant IgG level correlation (β): 2.9 (95%CI 0.7-5.1) |
| Theiler *et al.*, | NR | NR | **Maternal and delivery outcome N (%) (vaccinated vs** |
| 2021<sup>19</sup> |  |  | **unvaccinated)**<br>Adverse outcomes index (AOI): 91 (5%) vs 7 (5%); p= 0.9524<br>AOI excluding laceration: 55 (3%) vs 5 (4%); p= 0.6071<br>Hypoxic, ischaemic encephalopathy: 1 (0%) vs 0 (0%); p= 1<br>Uterine rupture, AOI: 1 (0%) vs 0 (0%); p= 1<br>Unplanned ICU admission: 2 (0%) vs 1 (1%); p= 0.1956<br>Birth trauma: 11 (1%) vs 0 (0%); p= 1<br>Return to OR: 6 (0%) vs 1 (1%); p= 0.3985<br>NICU admit > 2500g: 11 (1%) vs 1 (1%); p= 0.5821<br>5 Minute apgar <7: 38 (2%) vs 3 (2%); p= 0.7617<br>Hemorrhage with transfusion: 5 (0%) vs 1 (1%); p= 0.3531<br>Third- or fourth-degree laceration: 37 (2%) vs 2 (1%); p= 1<br>Mode of delivery: p= 0.6517<br>- spontaneous vaginal: 1238 (66%) vs 89 (64%)<br>- operative vaginal: 69 (4%) vs 7 (5%)<br>- cesarean: 555 (30%) vs 44 (31%)<br>Gestational age delivery: p=0.7028<br>- 37+: 1703 (91%) vs 127 (91%)<br>- 32-36.9: 134 (7%) vs 10 (7%)<br>- 24-31.9: 21 (1%) vs 2 (1%)<br>- <24: 4 (0%) vs 1 (1%)<br>Quantitative blood loss > 1000mL: 56 (3%) vs 6 (4%); p= 0.4452<br>Transfusion: 241 (13%) vs 25 (18%); p= 0.1198<br>Thromboembolism: 2 (0%) vs 0 (0%); p= 1<br>Stroke: 1 (0%) vs 0 (0%); p = 1<br>Eclampsia/pre-eclampsia (+/-72 h. of delivery): 23 (1%) vs 1(1%); p=1<br>Gestational hypertension: 225 (12%) vs 19 (14%); p= 0.6038<br>Low birth weight (<2500g): 121 (6%) vs 11 (8%); p= 0.5321<br>Very low birth weight (<1500g): 21 (1%) vs 3 (2%); p= 0.2332 |
|  |  |  | StillBirth: 6 (0%) vs 0 (0%); p= 1 |
| Beharier *et al.*, 2021<sup>20</sup> | NR | NR | **Temporal dependence in pregnant women**<br>After infection:<br>A gradual rise in IgG humoral response (Anti- S1, S2, RBD and N) was detected during the first 45 days after infection.<br>After the first dose:<br>In the same period, vaccinated participants receiving the first BNT162b2 dose showed a rapid IgG response to S1, S2, RBD but not N, resulting in high titer values by day 15 after the first dose.<br>After the second dose:<br>A further rise in IgG was observed following the second dose.<br>**Temporal dependence in neonates**<br>After the first dose:<br>The temporal dependence of fetal IgG for S1, S2 and RBD after vaccination trailed after the maternal IgG showing a significant response already by day 15.<br>After the second dose:<br>A further increase was observed following the second vaccination dose.<br>**Maternal IgG between vaccinated vs PCR-positive**<br>S1 IgG: higher in vaccination (P=0.0009)<br>RBD IgG: higher in vaccination (P=0.0045)<br>S2 IgG: higher in PCR-positive (P=0.016)<br>N IgG: higher in PCR-positive (P=<0.0001)<br>**Maternal to fetal IgG transfer ratio for S1, S2, RBD, and N**<br>PCR-positive vs N<sup>-</sup> group: Significant differences were found for S1, S2 and RBD (P<0.0002)<br>PCR-positive vs N<sup>+</sup> group: For all antibodies did not differ (P=0.4577)<br>**Maternal-fetal IgG response to infection and vaccination**<br>**correlation**<br>S1 IgG: R<sup>2</sup> = 0.9443; Adjusted R<sup>2</sup> = 0.9438; P<0.0001<br>S2 IgG: R<sup>2</sup> = 0.9353; Adjusted R<sup>2</sup> = 0.9348; P<0.0001<br>RBD IgG: R<sup>2</sup> = 0.9200; Adjusted R<sup>2</sup> = 0.9194; P<0.0001<br>N IgG: R<sup>2</sup> = 0.9366; Adjusted R<sup>2</sup> = 0.9361; P<0.0001<br>Infection vs vaccination maternal-fetal IgG response<br>- S1 IgG: P=0.2936<br>- S2 IgG: P=0.4212<br>- RBD IgG: P=0.0702<br>- N IgG: P=0.7616 |
| Paul and Chad, 2021<sup>21</sup> | NR | NR | **Cord blood Antibody**<br>IgG concentration: 1.31 U/mL |

Abbreviations: NR (not reported); AU (arbitrary unit)
\* It reached statistical significance (p=0.0369). However, the participant with a report of seizure has a known history of seizure disorder and her anticonvulsant blood level
was reported as borderline low.

### Infection Rate

Among pregnant women who received the Pfizer–BioNTech vaccine, 0.1% (3/2136) of pregnant women experienced Covid-19 infection within 14 days from vaccination and 0.4% (9/2136) after 14 days from vaccination. Additionally, among pregnant women who received Moderna vaccines, 0.4% (7/1822) of pregnant women experienced Covid-19 infection within 14 days from vaccination and 0.2% (3/1822) after 14 days from vaccination.^10^ Among unvaccinated vs vaccinated pregnant women, vaccination significantly reduced the risk of future infection (p=0.0004) and all infection cases, reported in the trimester I of vaccinated women, occurred prior to the first vaccination dose.^19^

### Maternal antibody response

Vaccination induced IgG and IgM production in 71% (87/122) pregnant women; 16% (19/122) pregnant women produced only IgG whilst 13% (16/122) had neither detectable IgG nor IgM.^14^ Vaccination provided a rapid immunologic response after the first dose while infection provided a gradual immunologic response. Moreover, administration of the second dose can further increase the IgG level among vaccinated women.^11,20^ Spike- and RBD-IgG titer rose rapidly after prime dose (p<0.0001 and p<0.01, respectively); and after receiving booster, it became higher than that of the prime dose (p<0.05 and p<0.001, respectively).^11^

Vaccination elicited IgG responses against S1, S2, and RBD but not N protein. Meanwhile, infection elicited all IgG responses against S1, S2, RBD, and N protein. Among vaccinated pregnant women, S1 IgG and RBD IgG levels were higher (p=0.0009 and p=0.0045, respectively). Yet, S2 IgG and N IgG were higher in infected pregnant women (p=0.016 and p<0.0001, respectively).^20^ Meanwhile, Gray et al., (2021) reported that Spike IgG titer is higher in vaccination than that of natural infection in pregnant women.^11^

Maternal SARS-CoV-2 spike protein IgG levels between vaccinated and uninfected unvaccinated pregnant women are 22.8±14.5 AU and 0.04±0.05 AU, respectively (p<0.001). While for the IgM levels are 4.1±13.2 AU and 0.19±0.12 AU, respectively (p=0.001).^13^ Among pregnant women that received two vaccine doses, anti-spike-protein IgG concentration median is 319 (211-1033) AU/mL and anti-RBD-Specific IgG concentration median is 11150 (6154-17575) AU/mL.^17^ Meanwhile, in two women that only received one dose of vaccine, the anti-spike-protein IgG concentrations are 50 AU/mL and 52 AU/m; and the anti-RBD-Specific IgG concentrations are 293 AU/mL and 1137 AU/mL.^17^

Antibody response in pregnant and non-pregnant women was evaluated by Collier et al., (2021).^12^ The study reported the median of IgG levels between vaccinated and infected pregnant women. The RBD IgG titers were 27601 and 1321, respectively; while the neutralizing antibody titers were 910 and 148, respectively. The median of RBD IgG titers between vaccinated and infected non-pregnant women were 37839 and 771, respectively; while for the Neutralizing antibody were 901 and 193, respectively.^12^

### Antibody transfer

A prospective case series reported that the IgG was detected in 89% (25/28) of cord blood, but none had detectable IgM.^18^ Moreover, antibody against SARS-CoV-2 RBD and neutralizing antibody was observed in cord blood. On vaccinated women, the maternal vs cord blood RBD IgG was 14953 AU vs 19873 AU, whilst for the neutralizing antibody was 1016 AU vs 324 AU.^12^ IgG against S protein was also detected in cord blood with concentration of 193 (111-260) AU/mL and its transfer ratio was 0.44 (0.25-0.61). Furthermore, the concentration of IgG against RBD was 3494 (1817-6163) AU/mL and its transfer ratio was 0.34 (0.27-0.56).^17^

Two different case report studies from mother who received two doses of BNT162b2 vaccine and mother who received one dose of Moderna vaccine, respectively, reported that SARS-CoV-2 specific IgG was detected in maternal blood and cord blood at a titer 1:25.600, while cord blood IgG concentration were detected at level of 1.31 U/mL.^15,21^

Regarding the number of doses received, antibody was detected in 98.5% (65/67) neonates from whom the mother received two doses of vaccine. From mothers who received only one vaccine dose, only 43.6% (24/55) neonates whose antibody was detected.^14^ Having received the second vaccine dose was positively correlated with infant IgG level (β=19.0 (95%CI 7.1-30.8)).^18^ In addition to doses, the interval from vaccination to delivery was correlated with IgG transfer ratio and infant IgG level. Increased latency from vaccination to delivery was positively correlated with IgG transfer ratio (β=0.2 (95%CI 0.1-0.2)) and infant IgG level (β=2.9 (95%CI 0.7-5.1)).^18^ For maternal-fetal IgG response, there was no statistical difference between vaccination and SARS-CoV-2 infection for S1 IgG (p=0.2936), S2 IgG (p=0.4212), RBD IgG (p=0.0702), and N IgG (p=0.7616).^20^

### Local adverse events

Among pregnant women, injection-site pain is the most common adverse event in both the Pfizer– BioNTech and Moderna vaccines. Following the Pfizer–BioNTech vaccination, as many as 84% (7602/9052) in the first dose and 89% (5886/6638) in the second dose experienced injection-site pain. While for the Moderna vaccine, 93% (7360/7930) and 96% (5388/5635) following the first and the second dose, respectively, experienced injection-site pain.^10^ It was also reported 88% (73/84) pregnant women experienced injection-site soreness following the first dose vaccination and 57% (44/84) following the second dose. Additionally, 75% (12/75) non-pregnant women experienced injection-site soreness after the first and the second dose of vaccine.^11^

Between pregnant and non-pregnant women, sore arms or pain were observed in 97% (37/38) pregnant women and 90% (894/991) non-pregnant women following Pfizer–BioNTech and Moderna vaccination.^16^

### Systemic adverse event

Systemic adverse events between two doses of Pfizer–BioNTech and Moderna were reported. Following Pfizer–BioNTech the first vs the second dose of vaccine, six most experienced systemic adverse events were fatigue (27% (2406/9052) vs 64% (4231/6638)); headache (17% (1497/9052) vs 47% (3138/6638)), myalgia (9% (795/9052) vs 44% (2916/6638)), chills (3% (254/9052) vs 26% (1747/6638)), fever (3% (256/9052) vs 25% (1648/6638)), and nausea (5% (492/9052) vs 20% (1356/6638)). For the Moderna vaccine, fatigue (33% (2616/7930) vs 81% (4541/5635)), headache (20% (1581/7930) vs 65% (3662/5635)), myalgia (15% (1167/7930) vs 66% (3722/5635)), chills (6% (442/7930) vs 49% (2755/5635)), fever (6% (453/7930) vs 46% (2594/5635)), and nausea (8% (638/7930) vs 34% (1909/5635)). Numerically, the incidence of each event is higher in the second dose. Moreover, the Moderna vaccine had more systemic adverse events than that of the Pfizer– BioNTech vaccine.^10^ Seizure was reported in a woman who received mRNA vaccine (p=0.0369), but it was known that the patient has a history of seizure disorder and the anticonvulsant level in the blood was borderline low.^16^

### Maternal outcomes

Maternal outcomes were described as pregnancy outcomes and delivery outcomes. Compared to unvaccinated pregnancy, vaccination did not significantly affect pregnancy or delivery outcomes. The adverse outcome index between vaccinated and unvaccinated pregnancy was insignificant (p=0.9524). Between these group, there were no statistical difference in pregnancy outcome such as eclampsia/pre-eclampsia (p=1), gestational hypertension (p=0.6038), gestational age (p=0.7028), and thromboembolism incidence (p=1).^19^ Moreover, the abortion rate was reported at 12.6% (104/827) and 9.4% (60/636) had preterm birth. Statistically, vaccination also did not affect delivery outcomes such as birth trauma (p=1), uterine rupture (p=1), unplanned ICU admission (p=0.1956), quantitative blood loss>1000mL (p=0.4452) or hemorrhage with transfusion (p=0.3531), mode of delivery (p=0.6517), and stillbirth (p=1).^19^

### Neonatal outcomes

As many as 15% (2/13) required NICU admission, 8% (1/13) experienced TTN, and 8% (1/13) required supplemental oxygen or CPAP. Preterm delivery was reported in 8% (1/13) of women.^11^ Moreover, 0.1% (1/725) were stillbirth, 3.2% (23/724) had small size for gestational age and 2.2% (16/724) had congenital anomalies. No neonatal death was reported.^10^ On statistical analysis, NICU admission (p=0.5821), 5 minute Apgar <7 (0.7617), hypoxic ischemic encephalopathy (p=1), and low birth weight (p= 0.5321) or very low birth weight (p= 0.2332), did not differ significantly between vaccinated and unvaccinated women.^19^

## DISCUSSION

### Main Finding

It has been described that Pfizer-BioNTech and Moderna vaccines were efficacious for preventing future SARS-CoV-2 infection in pregnant women.^19^ Following Pfizer-BioNTech and Moderna vaccinations, the vast majority of pregnant women will have injection-site pain or soreness.^10,11,16^ Some most-experienced systemic adverse events were fatigue, headache, chills, myalgia, fever, and nausea.^10,16^ The incidence of these systemic adverse events were higher after the second dose compared to the first dose.^10-12^ Numerically, there were more individuals who experienced these systemic adverse events in Moderna vaccine group than that of Pfizer-BioNTech group.^10^ The adverse events were not statistically differ between pregnant and non pregnant women, except seizure. However, the woman was known to have a history of seizure disorder and the anticonvulsant level was measured borderline-low.^19^ Interestingly, the pregnancy, delivery, and neonatal outcomes did not differ between pregnant and non pregnant women.^19^

Maternal antibody response has been described formed following vaccination.^11-15,17,18,20^ Through vaccination, antibody response was formed rapidly; while through vaccination it was formed gradually.^20^ The number of IgG and IgM against SARS-CoV-2 were significantly increased after vaccination. The response was increased after a booster was given.^11,13^ Although vast majority of pregnant women had IgG seroconversion, IgM seroconversions were observed in a minority of pregnant women.^18^ After vaccination, IgG against S1, S2 and RBD formed whilst IgG against S1, S2, RBD and N protein formed following natural infections. Moreover, S1- and RBD-IgG were observed higher in vaccinated pregnant women. Meanwhile the S2- and N-IgG were observed higher in naturally infected pregnant women.^20^ Also, the RBD IgG and neutralizing antibody was higher in vaccinated individuals than that of naturally infected.^12^

Antibody transfer was also reported.^12,14,15,17,18,20,21^ Cord blood antibody and maternal antibody levels were about equal.^18^ Additionally, latency and number of doses were correlated with the antibody transfer strength.^14,18^ The longer the latency, the better the antibody transfer and the higher the IgG.^18^ A mother who received two doses of vaccines would have her infant getting higher IgG levels.^18^ Between infection and vaccination, there was no difference in maternal-fetal IgG response.^20^

### Finding on other studies

A sufficient herd immunity threshold in a population will provide indirect protection for susceptible individuals from infected hosts.^22^ The threshold varies across different infections.^23-25^ In Covid-19, it was said that for a vaccine with 100% efficacy and providing life-long protection, we would need 60-75% herd immunity. This required number would be increased if the vaccine efficacy was reduced to 85%; a 75-90% herd immunity would be required.^26^ Although we described that Pfizer-BioNTech and Moderna vaccines are efficacious to prevent future SARS-CoV-2 infection, whether the efficacy of these vaccines is reduced in pregnancy remains unknown.

This herd immunity threshold may also vary across populations. It depends on some factors--epidemiological factors (e.g., population density and transmission dynamics) and immunological factors (e.g., immune stats of population).^22^ An approach to achieve this threshold is through mass vaccination campaigns.^22^ Thus, high vaccination coverage is important for achieving sufficient herd immunity.^27,28^

On note, not only pregnant individuals but also those who were not pregnant will experience these adverse reactions as well. In Moderna and Pfizer-BioNTech studies where pregnant women and children were excluded, the incidence of systemic adverse events increased after the second dose of vaccine.^29,30^ Following the Moderna vaccine booster, local pain was experienced by 83.2-89.9% of individuals, followed by fatigue (58.3-65.3%) and headache (46.2-62.8%). Meanwhile, there were 10.0-15.5% who experienced fever. These were generally resolved within 3.1-3.2 days.^29^ Following the Pfizer-BioNTech vaccine booster, injection-site pain was experienced by 66%-78% individuals, followed by fatigue (51-59%) and headache (39-52%). Meanwhile, fever was experienced by 11-16% of individuals. These adverse events were transient, resolved within 1-2 days.^30^

Safety is an important concern for Covid-19 vaccination in pregnant women. Based on a survey in 16 countries, pregnant women were less likely to accept vaccines for themselves. Given that a Covid-19 vaccine had 90% efficacy, 73.4% non-pregnant women intended to receive the vaccine while pregnant women were only 52%. The confidence in vaccine safety and effectiveness was one of the predictors for vaccine acceptance.^31^ However, vaccination safety did not differ among those who are pregnant or not. Moreover, vaccination did not affect pregnancy, delivery, and pregnancy outcomes. On the other hand, some conditions during pregnancy made pregnant women susceptible to severe disease of Covid-19.^32^

During pregnancy, the upper respiratory tract was swollen and lung expansion was restricted. It makes pregnant women more susceptible to respiratory pathogens.^33^ Moreover, pro-inflammatory phase was present in the first trimester, for the implantation of the embryo and placenta, and third trimester, to prepare for the initiation of labor.^34^ In fact, severe Covid-19 is associated with cytokine storms.^35^ The pro-inflammatory phase during the first and third trimester of pregnancy, made them to be more prone to severe Covid-19.^36^

Individuals with significant medical comorbidities, the elderly, and pregnant women are highly vulnerable to Covid-19 infection.^37,38^ Although newborns and infants are less likely to get SARS-CoV-2 infections, they are more prone to severe SARS-CoV-2 infection.^37,39,40^ Passive immunity in neonates may be potentially protective. Administration of booster doses can enhance antibody transfer which may provide a better immunity in neonates. However, this fetal passive immunity may be altered due to placental sieving^41^, depending on the gestational age at first vaccine dose or infection. The longer latency interval from first dose of vaccination to delivery should be required for better antibody transfer.^18^

In contrast to natural viral infection such as in Zika dan DENV,^42,43^ antibody transfer observed following other vaccinations such as in pertussis and Influenza vaccines.^44,45,46^ Timeframe following natural SARS-CoV-2 infection may affect antibody transfer. An earlier Covid-19 infection may provide a better placental antibody transfer.^20^ Additionally, poor placental antibody transfer was exclusively observed only in the third trimester of pregnancy, even though the maternal antibody response was significantly higher.^47^ Natural infection may result in higher morbidity and mortality^48^, so vaccination should be the better option for the dyads.

Many SARS-CoV-2 mutations had altered Covid-19 transmissibility and, probably, severity.^49,50,51^ Moreover, these mutations can also impact a treatment efficacy, especially monoclonal antibody treatments due to immune escape.^52,53,54^ It had been reported, compared to the wild type of USA-WA1/2020 SARS-CoV-2, the binding strength of neutralizing antibodies formed after Pfizer and Moderna vaccination was observed 3.5- and 6-fold lower for B.1.1.7 and B.1351, respectively.^12^ Antibody resistance against B.1.1.7 and B.1351 was also reported by another study.^55^ In addition, in UK, it also was reported that B.1.617.2 had overtaken B.1.1.7. variants.^56^ Maximizing the second dose vaccine coverage may provide a stronger protection against this variant.^57^ However, we still need to know whether we would need a new vaccine or booster dose since there are evidence of reduced efficacy of currently available vaccine for these strains. This problem should be taken more seriously, and other public health measures are necessary as an effort to end this pandemic.^58^

### Strength and limitation

To the best of our knowledge, this systematic review used the most recent evidence to describe the efficacy, safety and immunogenicity of Covid-19 mRNA vaccine in pregnancy. All studies included in this review were assessed as high quality studies. However, studies are all observational studies due to no RCTs reports of Covid-19 vaccination for pregnant women yet currently available. These studies reported only from mRNA type vaccines. Moreover, all available studies that were included were only from the United States and Israel.

## CONCLUSION

In this study, mRNA vaccines, especially Pfizer-BioNTech and Moderna vaccines, are efficacious for preventing future SARS-CoV-2 infections. These vaccines can induce antibody responses for pregnant women and their fetus. Pregnant women should be given two doses of vaccine for more robust maternal and fetal antibody response. Longer latency was associated with more robust fetal antibody response. Almost all pregnant women who received vaccination, either in the first or second dose, will experience injection-site pain. Furthermore, the second dose of vaccine will produce more systemic adverse events than that of the first dose and administration of Moderna vaccine was observed to have a more frequent systemic adverse events. Biologically speaking, we may conclude that for a short term, vaccination did not affect pregnancy, delivery, or neonatal outcomes.

## Data Availability

Details of data availability are available from the first author on request

## Data Availability

Details of data availability are available from the first author on request

## Data Availability

Details of data availability are available from the first author on request

## REFERENCES

1. Dashraath P, Wong JLJ, Lim MXK, Lim LM, Li S, Biswas A, et al. Coronavirus disease 2019 (COVID-19) pandemic and pregnancy. Am J Obstet Gynecol 2020;222(6):521–531. doi: 10.1016/j.ajog.2020.03.021.

2. Huang C, Wang Y, Li X, Ren L, Zhao J, Hu Y, et al. Clinical features of patients infected with 2019 novel coronavirus in Wuhan, China. The Lancet 2020;395(10223):497–506. doi: 10.1016/S0140-6736(20)30183-5.

3. Zambrano LD, Ellington S, Strid P, Galang RR, Oduyebo T, Tong VT, et al. Update: Characteristics of Symptomatic Women of Reproductive Age with Laboratory-Confirmed SARS-CoV-2 Infection by Pregnancy Status - United States, January 22-October 3, 2020. MMWR Morb Mortal Wkly Rep 2020;69(44):1641–1647. doi:10.15585/mmwr.mm6944e3

4. Gray KJ, Bordt EA, Atyeo C, Deriso E, Akinwunmi B, Young N, et al. Coronavirus disease 2019 vaccine response in pregnant and lactating women: a cohort study. Am J Obstet Gynecol 2021;26:S0002-9378(21)00187-3. doi: 10.1016/j.ajog.2021.03.023.

5. Sutton D, Fuchs K, D’Alton M, Goffman D. Universal Screening for SARS-CoV-2 in Women Admitted for Delivery. N Engl J Med. 2020;28;382(22):2163–2164. doi: 10.1056/NEJMc2009316

6. Yanes-Lane M, Winters N, Fregonese F, Bastos M, Perlman-Arrow S, Campbell JR, et al. Proportion of asymptomatic infection among COVID-19 positive persons and their transmission potential: A systematic review and meta-analysis. PLoS One 2020;3;15(11):e0241536. doi: 10.1371/journal.pone.0241536.

7. Page MJ, McKenzie JE, Bossuyt PM, Boutron I, Hoffmann TC, Mulrow CD, et al. The PRISMA 2020 statement: an updated guideline for reporting systematic reviews. BMJ 2021;372:71. doi:10.1136/bmj.n71

8. Wells G, Shea B, O’Connell D, Peterson J, Welch V, Losos M, et al. The Newcastle-Ottawa Scale (NOS) for assessing the quality of nonrandomised studies in meta-analyses. Accessed June 27, 2021. http://www.ohri.ca/programs/clinical_epidemiology/oxford.asp

9. Joanna Briggs Institute. Checklist for systematic reviews and research syntheses. Accessed June 26, 2021. https://jbi.global/critical-appraisal-tools

10. Shimabukuro TT, Kim SY, Myers TR, Podo PL, Oduyebo T, Panagiotakopoulos L, et al. Preliminary Findings of mRNA Covid-19 Vaccine Safety in Pregnant Persons. N Engl J Med 2021;384(24):2273–2282. doi:10.1056/NEJMoa2104983

11. Gray KJ, Bordt EA, Atyeo C, Deriso E, Akinwunmi B, Young N, et al. Coronavirus disease 2019 vaccine response in pregnant and lactating women: a cohort study. Am J Obstet Gynecol, 2021;S0002-9378(21)00187-3. doi:10.1016/j.ajog.2021.03.023

12. Collier AY, McMahan K, Yu J, Tostanoski LH, Aguayo R, Ansel J, et al. Immunogenicity of COVID-19 mRNA Vaccines in Pregnant and Lactating Women. JAMA 2021;325(23):2370–2380. doi:10.1001/jama.2021.7563

13. Shanes ED, Otero S, Mithal LB, Mupanomunda CA, Miller ES, Goldstein JA. Severe Acute Respiratory Syndrome Coronavirus 2 (SARS-CoV-2) Vaccination in Pregnancy: Measures of Immunity and Placental Histopathology. Obstet Gynecol 2021;10.1097/AOG.0000000000004457. doi:10.1097/AOG.0000000000004457

14. Prabhu M, Murphy EA, Sukhu AC, Yee J, Singh S, Eng D, et al. Antibody response to SARS-CoV-2 mRNA vaccines in pregnant women and their neonates. bioRxiv (preprint) 2021;2021.04.05.438524.

15. Gill L, Jones CW. Severe Acute Respiratory Syndrome Coronavirus 2 (SARS-CoV-2) Antibodies in Neonatal Cord Blood After Vaccination in Pregnancy. Obstet Gynecol 2021;137(5):894–896. doi:10.1097/AOG.0000000000004367

16. Kadali RAK, Janagama R, Peruru SR, Racherla S, Tirumala R, Madathala RR, et al. Adverse effects of COVID-19 messenger RNA vaccines among pregnant women: a cross-sectional study on healthcare workers with detailed self-reported symptoms. Am J Obstet Gynecol 2021;10:S0002-9378(21)00638-4. doi:10.1016/j.ajog.2021.06.007

17. Rottenstreich A, Zarbiv G, Oiknine-Djian E, Zigron R, Wolf DG, Porat S. Efficient maternofetal transplacental transfer of anti-SARS-CoV-2 spike antibodies after antenatal SARS-CoV-2 BNT162b2 mRNA vaccination. Clin Infect Dis 2021;3:ciab266. doi:10.1093/cid/ciab266

18. Mithal LB, Otero S, Shanes ED, Goldstein JA, Miller ES. Cord blood antibodies following maternal coronavirus disease 2019 vaccination during pregnancy. Am J Obstet Gynecol 2021;1:S0002-9378(21)00215-5. doi:10.1016/j.ajog.2021.03.035

19. Theiler RN, Wick M, Mehta R, Weaver A, Virk A, Swift M. Pregnancy and birth outcomes after SARS-CoV-2 vaccination in pregnancy. medRxiv (preprint) 2021;2021.05.17.21257337.

20. Beharier O, Plitman Mayo R, Raz T, Nahum Sacks K, Schreiber L, Suissa-Cohen Y, et al. Efficient maternal to neonatal transfer of antibodies against SARS-CoV-2 and BNT162b2 mRNA COVID-19 vaccine. J Clin Invest 2021;150319. doi:10.1172/JCI150319

21. Paul G, Chad R. Newborn antibodies to SARS-CoV-2 detected in cord blood after maternal vaccination – a case report. BMC Pediatr 2021;21:138. doi: 10.1186/s12887-021-02618-y

22. Randolph H, Barreiro L. Herd Immunity: Understanding COVID-19. Immunity 2020;52(5):737–741. doi:10.1016/j.immuni.2020.04.012

23. Vignesh R, Shankar E, Velu V, Thyagarajan S. Is Herd Immunity Against SARS-CoV-2 a Silver Lining ?. Frontiers In Immunology 2020;11:586781. doi: 10.3389/fimmu.2020.586781

24. Xia Y, Zhong L, Tan J, Zhang Z, Lyu J, Chen Y, et al. How to Understand “Herd Immunity” in COVID-19 Pandemic. Frontiers In Cell And Developmental Biology 2020;8:547314. doi: 10.3389/fcell.2020.547314

25. Griffith B, Ulrich A, Becker A, Nederhoff D, Koch B, Awan F, et al. Does education about local vaccination rates and the importance of herd immunity change US parents’ concern about measles?. Vaccine 2020;38(50), 8040–8048. doi:10.1016/j.vaccine.2020.09.076

26. Anderson R, Vegvari C, Truscott J, Collyer B. Challenges in creating herd immunity to SARS-CoV-2 infection by mass vaccination. The Lancet 2020;396(10263):1614–1616. doi: 10.1016/S0140-6736(20)32318-7

27. Lahariya C. Vaccine epidemiology: A review. Journal Of Family Medicine And Primary Care 2016;5(1):7. doi: 10.4103/2249-4863.184616

28. Orenstein W, Seib K. Mounting a Good Offense against Measles. New England Journal Of Medicine 2014;371(18):1661–1663. doi: 10.1056/NEJMp1408696

29. Baden LR, El Sahly HM, Essink B, Kotloff K, Frey S, Novak R, et al. Efficacy and Safety of the mRNA-1273 SARS-CoV-2 Vaccine. New England Journal Of Medicine 2021;384(5):403–416. doi: 10.1056/NEJMoa2035389

30. Polack F, Thomas S, Kitchin N, Absalon J, Gurtman A, Lockhart S, et al. Safety and Efficacy of the BNT162b2 mRNA Covid-19 Vaccine. New England Journal Of Medicine 2020;383(27):2603–2615. doi:10.1056/NEJMoa2034577

31. Skjefte M, Ngirbabul M, Akeju O, Escudero D, Hernandez-Diaz S, Wyszynski D, et al. COVID-19 vaccine acceptance among pregnant women and mothers of young children: results of a survey in 16 countries. European Journal Of Epidemiology 2021;36(2):197–211. doi:10.1007/s10654-021-00728-6

32. Allotey J, Stallings E, Bonet M, Yap M, Chatterjee S, Kew T, et al. Clinical manifestations, risk factors, and maternal and perinatal outcomes of coronavirus disease 2019 in pregnancy: living systematic review and meta-analysis. BMJ 2020;370:m3320. doi: 10.1136/bmj.m3320

33. Cavalcante MB, Cavalcante CTMB, Sarno M, Barini R, Kwak-Kim J. Maternal immune responses and obstetrical outcomes of pregnant women with COVID-19 and possible health risks of offspring. J Reprod Immunol 2021;2;143:103250. doi: 10.1016/j.jri.2020.103250

34. Mor G. The unique immunological and microbial aspects of pregnancy. Nat. Rev. Immunol 2017;17:469–482. doi: 10.1038/nri.2017.64

35. Huang C. Clinical features of patients infected with 2019 novel coronavirus in Wuhan, China. Lancet 2020;395(10223):497–506. doi: 10.1016/S0140-6736(20)30183-5

36. Zambrano LD, Ellington S, Strid P, Galang RR, Oduyebo T, Tong VT, et al. Update: Characteristics of Symptomatic Women of Reproductive Age with Laboratory-Confirmed SARS-CoV-2 Infection by Pregnancy Status - United States, January 22-October 3, 2020. MMWR Morb Mortal Wkly Rep 2020;6;69(44):1641–1647. doi: 10.15585/mmwr.mm6944e3.

37. Levin A, Hanage W, Owusu-Boaitey N, Cochran K, Walsh S, Meyerowitz-Katz G. Assessing the age specificity of infection fatality rates for COVID-19: systematic review, meta-analysis, and public policy implications. European Journal Of Epidemiology 2020;35(12), 1123–1138. doi: 10.1007/s10654-020-00698-1

38. Zheng Z, Peng F, Xu B, Zhao J, Liu H, Peng J, et al. Risk factors of critical & mortal COVID-19 cases: A systematic literature review and meta-analysis. Journal Of Infection 2020;81(2):e16–e25. doi: 10.1016/j.jinf.2020.04.021.

39. Liguoro I, Pilotto C, Bonanni M, Ferrari M, Pusiol A, Nocerino A, et al. SARS-COV-2 infection in children and newborns: a systematic review. European Journal Of Pediatrics 2020;179(7):1029–1046. doi: 10.1007/s00431-020-03684-7

40. Dong Y, Mo X, Hu Y, Qi X, Jiang F, Jiang Z, et al. Epidemiology of COVID-19 Among Children in China. Pediatrics 2020;145(6):e20200702. doi:10.1542/peds.2020-0702

41. Jennewein M, Goldfarb I, Dolatshahi S, Cosgrove C, Noelette F, Krykbaeva M, et al. Fc Glycan-Mediated Regulation of Placental Antibody Transfer. Cell 2019;178(1), 202-215.e14. doi: 10.1016/j.cell.2019.05.044

42. Atyeo C, Pullen K, Bordt E, Fischinger S, Burke J, Michell A, et al. Compromised SARS-CoV-2-specific placental antibody transfer. Cell 2021;184(3):628-642.e10. doi: 10.1016/j.cell.2020.12.027

43. Haghpanah F, Lin G, Levin S, Klein E. Analysis of the potential impact of durability, timing, and transmission blocking of COVID-19 vaccine on morbidity and mortality. Eclinicalmedicine 2021;35;100863. doi: 10.1016/j.eclinm.2021.100863

44. Castanha PMS, Souza WV, Braga C, Araújo Tvb, Ximenes RAA, Albuquerque Mfpm, et al. Perinatal analyses of Zika- and dengue virus-specific neutralizing antibodies: A microcephaly case-control study in an area of high dengue endemicity in Brazil. PLoS Negl Trop Dis 2019;11;13(3):e0007246. doi: 10.1371/journal.pntd.0007246.

45. Perret C, Chanthavanich P, Pengsaa K, Limkittikul K, Hutajaroen P, Bunn JE, et al. Dengue infection during pregnancy and transplacental antibody transfer in Thai mothers. J Infect 2005;51(4):287–93. doi: 10.1016/j.jinf.2004.10.003.

46. Gonçalves G, Cutts FT, Hills M, Rebelo-Andrade H, Trigo FA, Barros H. Transplacental transfer of measles and total IgG. Epidemiol Infect 1999;122(2):273–9. doi: 10.1017/s0950268899002046.

47. Heininger U, Riffelmann M, Leineweber B, Wirsing von Koenig CH. Maternally derived antibodies against Bordetella pertussis antigens pertussis toxin and filamentous hemagglutinin in preterm and full term newborns. Pediatr Infect Dis J 2009;28(5):443–5. doi: 10.1097/INF.0b013e318193ead7.

48. Munoz FM, Bond NH, Maccato M, Pinell P, Hammill HA, Swamy GK, et al. Safety and immunogenicity of tetanus diphtheria and acellular pertussis (Tdap) immunization during pregnancy in mothers and infants: a randomized clinical trial. JAMA 2014;7;311(17):1760–9. doi: 10.1001/jama.2014.3633.

49. Korber B, Fischer W, Gnanakaran S, Yoon H, Theiler J, Abfalterer W et al. Tracking Changes in SARS-CoV-2 Spike: Evidence that D614G Increases Infectivity of the COVID-19 Virus. Cell. 2020;182(4):812-827.e19. doi: 10.1016/j.cell.2020.06.043

50. Bian L, Gao F, Zhang J, He Q, Mao Q, Xu M et al. Effects of SARS-CoV-2 variants on vaccine efficacy and response strategies. Expert Review of Vaccines. 2021;:1–9. doi: 10.1080/14760584.2021.1903879

51. Gómez C, Perdiguero B, Esteban M. Emerging SARS-CoV-2 Variants and Impact in Global Vaccination Programs against SARS-CoV-2/COVID-19. Vaccines. 2021;9(3):243. doi: 10.3390/vaccines9030243

52. Boehm E, Kronig I, Neher R, Eckerle I, Vetter P, Kaiser L. Novel SARS-CoV-2 variants: the pandemics within the pandemic. Clinical Microbiology and Infection. 2021. doi: 10.1016/j.cmi.2021.05.022

53. Hoffmann M, Arora P, Groß R, Seidel A, Hörnich B, Hahn A et al. SARS-CoV-2 variants B.1.351 and P.1 escape from neutralizing antibodies. Cell. 2021;184(9):2384-2393.e12. doi: 10.1016/j.cell.2021.03.036. doi:10.1016/j.cell.2021.03.036

54. Starr T, Greaney A, Dingens A, Bloom J. Complete map of SARS-CoV-2 RBD mutations that escape the monoclonal antibody LY-CoV555 and its cocktail with LY-CoV016. Cell Reports Medicine. 2021;2(4):100255. doi: 10.1016/j.xcrm.2021.100255

55. Wang P, Nair M, Liu L, Iketani S, Luo Y, Guo Y et al. Antibody Resistance of SARS-CoV-2 Variants B.1.351 and B.1.1.7. 2021. doi: 10.1101/2021.01.25.428137

56. Torjesen I. Covid-19: Delta variant is now UK’s most dominant strain and spreading through schools. BMJ 2021;373:n1445. doi:10.1136/bmj.n144. doi:10.1136/bmj.n144

57. Bernal J, Andrews N, Gower C, Gallagher E, Simmons R, Thelwall S et al. Effectiveness of COVID-19 vaccines against the B.1.617.2 variant. 2021. doi: https://doi.org/10.1101/2021.05.22.21257658

58. Ayouni I, Maatoug J, Dhouib W, Zammit N, Fredj S, Ghammam R et al. Effective public health measures to mitigate the spread of COVID-19: a systematic review. BMC Public Health. 2021;21(1). doi: 10.1186/s12889-021-11111-1

