## Supplementary Materials for "Covid-19 Vaccination in Pregnancy: A Systematic Review"

**Table 1. Newcastle-Ottawa Scale (NOS) quality assessment of each included cohort study**

| Study | **Selection** | | | | **Comparability** | | **Outcome** | | |  |
| --- | --- | --- | --- | --- | --- | --- | --- | --- | --- | --- |
|  | Representativeness of exposed cohort | Selection of nonexposed cohort | Ascertainment of exposure | Demonstration that outcome of interest was not present at start of study | Adjust for the most important risk factors | Adjust for other risk factors | Assessment of outcome | Follow-up length | Loss to follow-up rate | Total quality score |
| Shimabukuro et al., 2021 | 🟊 | 🟊 | - | - | 🟊 | 🟊 | 🟊 | 🟊 | 🟊 | 7 |
| Gray et al., 2021 | 🟊 | 🟊 | 🟊 | - | 🟊 | 🟊 | 🟊 | 🟊 | 🟊 | 8 |
| Collier et al., 2021 | - | 🟊 | 🟊 | - | 🟊 | 🟊 | 🟊 | 🟊 | 🟊 | 7 |
| Shanes et al., 2021 | 🟊 | 🟊 | 🟊 | 🟊 | 🟊 | 🟊 | 🟊 | 🟊 | 🟊 | 9 |
| Prabhu et al., 2021 | - | - | 🟊 | 🟊 | 🟊 | 🟊 | 🟊 | 🟊 | 🟊 | 7 |
| Rottenstreich et al., 2021 | 🟊 | - | 🟊 | - | 🟊 | 🟊 | 🟊 | 🟊 | 🟊 | 7 |
| Theiler et al., 2021 | 🟊 | 🟊 | 🟊 | - | 🟊 | 🟊 | 🟊 | 🟊 | 🟊 | 8 |
| Beharier et al., 2021 | 🟊 | 🟊 | 🟊 | 🟊 | 🟊 | 🟊 | 🟊 | 🟊 | 🟊 | 9 |

**Table 2. Joanna Briggs Institute (JBI) critical appraisal for case series study**

| No | Checklist questions | Mithal et al., 2021 |
| --- | --- | --- |
| 1. | Were there clear criteria for inclusion in the case series? | Yes |
| 2. | Was the condition measured in a standard, reliable way for all participants included in the case series? | Yes |
| 3. | Were valid methods used for identification of the condition for all participants included in the case series? | Yes |
| 4. | Did the case series have consecutive inclusion of participants? | Yes |
| 5. | Did the case series have complete inclusion of participants? | Yes |
| 6. | Was there clear reporting of the demographics of the participants in the study? | Yes |
| 7. | Was there clear reporting of clinical information of the participants? | Yes |
| 8. | Were the outcomes or follow up results of cases clearly reported? | Yes |
| 9. | Was there clear reporting of the presenting site(s)/clinic(s) demographic information? | Yes |
| 10. | Was statistical analysis appropriate? | Yes |

**Table 3. Joanna Briggs Institute (JBI) critical appraisal for case report study**

| No | Checklist questions | Gill and Jones, 2021 | Paul and Chad, 2021 |
| --- | --- | --- | --- |
| 1. | Were patient’s demographic characteristics clearly described? | Yes | No |
| 2. | Was the patient’s history clearly described and presented as a timeline? | Yes | Yes |
| 3. | Was the current clinical condition of the patient on presentation clearly described? | Yes | Yes |
| 4. | Were diagnostic tests or assessment methods and the results clearly described? | Yes | Yes |
| 5. | Was the intervention(s) or treatment procedure(s) clearly described? | Yes | Yes |
| 6. | Was the post-intervention clinical condition clearly described? | Yes | Yes |
| 7. | Were adverse events (harms) or unanticipated events identified and described? | Yes | Unclear |
| 8. | Does the case report provide takeaway lessons? | Yes | Yes |

**Table 4. Joanna Briggs Institute (JBI) critical appraisal for cross-sectional study**

| No | Checklist questions | Kadali et al., 2021 |
| --- | --- | --- |
| 1. | Were the criteria for inclusion in the sample clearly defined? | Yes |
| 2. | Were the study subjects and the setting described in detail? | Unclear |
| 3. | Was the exposure measured in a valid and reliable way? | Yes |
| 4. | Were objective, standard criteria used for measurement of the condition? | Yes |
| 5. | Were confounding factors identified? | No |
| 6. | Were strategies to deal with confounding factors stated? | No |
| 7. | Were the outcomes measured in a valid and reliable way? | Yes |
| 8. | Was appropriate statistical analysis used? | Yes |
